# Identifying mental health service needs of people in Australian prisons

**DOI:** 10.64898/2026.02.18.26346585

**Authors:** Charlotte Comben, Meredith Burgess, Zoe Rutherford, Carla Meurk, Lorena Rivas, Julie John, Sandra Diminic

**Author notes:** **Corresponding author:** Charlotte Comben, School of Public Health, The University of Queensland, Brisbane, QLD, Australia. **Author contributions:** CC, ZR, CM and SD conceived the study. CC, JJ and SD designed and conducted focus groups and online survey. CC, MB, ZR, CM and LR analysed results. The manuscript was prepared by CC with critical input from ZR, CM and SD. All authors read and approved the final manuscript.

## Abstract

**Objective:** This study aimed to identify characteristics that define population need groups with similar mental health service needs within prisons and describe the mix of services required to meet those needs.

**Methods:** Mixed methods were used, including three iterative, semi-structured focus groups, followed by an online survey, seeking information on the characteristics that define service needs, how these can identify groups of people who require mental health care in prisons and the services required by each group. Participation was sought from prison health services, prison mental health services, non-government service partners and people with a lived experience. Focus group transcripts and free text survey responses were thematically analysed. Descriptive statistics were generated for online survey responses to Likert Scales to determine the levels of agreement with survey content.

**Results:** The characteristics and service needs of four distinct population groups who require mental health care in prisons were defined: indicated prevention, mild, moderate, severe and complex. These groups were delineated using characteristics including presence of a diagnosed mental illness, level of functional impairment, presence of added complexity and service response required. The required service mix varied across need groups, however service types common across all groups included assessments, psychological therapies, peer support, lifestyle interventions and carer support.

**Conclusions:** The identified need groups and service descriptions will contribute to the evidence required for needs-based planning of mental health care in Australian prisons. This information can be used for planning a responsive, equitable, and needs-based mental health service system within custodial environments.

## Introduction

Rates of recent and lifetime mental illness are notably higher in prisons than in the general Australian population (Australian Bureau of Statistics, 2022b; Comben et al., 2026; Yee et al., 2024). Some people in prison experience complex mental health needs alongside co-occurring physical health conditions, intellectual disability, and substance use disorders (Butler et al., 2007; Butler et al., 2011). For some, incarceration represents their first point of access to mental health care (Ogloff, 2002). This presents a critical public health opportunity to deliver care that may improve outcomes during and after incarceration.

Responding effectively to mental health care need in prisons requires needs-based health service planning to ensure services align to population needs. This is a priority, as people in Australian prisons are excluded from receiving Medicare, the country’s universal health care program and can only access mental health care via prison-based health services funded by state and territory governments (Linnane et al., 2023). Moreover, Australia does not have a nationally consistent model of mental health care service delivery in prisons, nor are there nationally consistent outcome measures or standards (Productivity Commission, 2020). Jurisdictions operate independent prison systems, with funding and service delivery reflective of differing political discourses, cultures, and legislative and historical contexts (Bartels et al., 2018; Productivity Commission, 2021). Similar challenges in planning mental health care in prisons exist outside of Australia; recent evidence suggests demand exceeds available resources in Scottish prisons as workforce is based on historical service use rather than current need (Gilling McIntosh et al., 2023).

Australia’s National Mental Health Service Planning Framework (NMHSPF) is a needs-based planning model that estimates services required to meet population mental healthcare needs but currently excludes justice-involved populations. Developing an evidence base to enable such modelling is essential, given the high levels of mental illness within Australian prisons and the vulnerabilities faced by these populations. A range of evidence is required to undertake such planning, including information about who needs mental health care in prisons and the types of mental health care they require (World Health Organisation, 2003).

In needs-based planning, service needs are multidimensional (Australian Government Department of Health, 2020; Diminic et al., 2023), with attributes including level of functional impairment, complexity, social and environmental stressors, and/or supports related to the presence of mental illness. These attributes can be used to delineate population need groups who have identifiable differences in service needs (Diminic et al., 2023). A mix of services can then be described for each population need group to model resourcing required to deliver such care. Needs-based approaches have demonstrated impacts on outcomes, including reduced odds of returning to prison (Long et al., 2018).

Population need groups should be based on care needs rather than diagnosis (Patel et al., 2023; Segal et al., 2018; Suetani et al., 2024). Diagnostic information alone is not a sufficient indicator of mental health service needs for workforce planning (Segal et al., 2018), as service responses and competencies required to deliver care are often diagnosis agnostic. Previous research has enumerated individual drivers of prison mental health service needs such as levels of mental illness/psychosocial distress (Comben et al., 2026; Yee et al., 2024), substance use (Butler et al., 2011; Holmwood et al., 2008; Kerslake et al., 2020; Richmond et al., 2013), intellectual disability (Trofimovs et al., 2021), and historical trauma (Egeressy et al., 2009; Goddard and Pooley, 2019). However, these drivers have previously not been examined together to understand holistic levels of need across individuals and groups.

Therefore, the aims of the current study were to (1) define need groups of people with similar mental health service needs in prisons based on the domains that drive service needs; and (2) describe the mix of mental health services required for each identified group. The need groups and service descriptions will contribute to the evidence for needs-based planning of mental health care in Australian prisons, and can be used for planning a responsive, equitable, and needs-based mental health service system within custodial environments.

## Methods

### Design

This study examined mental health service needs of those in prison, in the context of a larger study developing inputs for a needs-based forensic mental health service planning model for adults (18+ years) in Australia. A mixed-methods approach with a sequential design comprising three focus groups and an online survey was used (Figure 1). Ethics approval was obtained from the UQ Human Research Ethics Committee (2021/HE002377) and reporting adheres to the Good Reporting of a Mixed Methods Study guidance (O’Cathain et al., 2017).

**Figure 1.**
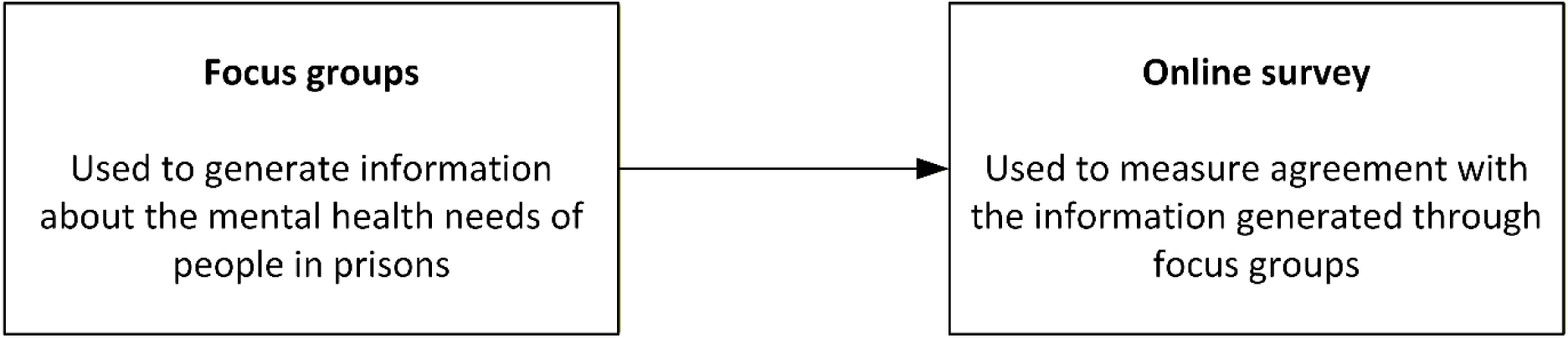
Sequential study design.

### Participants and recruitment

Recruitment for focus group participants focussed on three stakeholder groups responsible for the provision of mental health care in prisons: (1) prison/custodial/corrective health services; (2) prison mental health services; and (3) NGO service partners, with participation sought from service directors, service managers, and service providers. Additionally, people with a lived experience of mental illness in prison, or their carers, were invited.

Invitations were sent via email to a network of Australian senior forensic mental health clinicians and relevant organisations, such as professional bodies (e.g., Australian Psychological Society College of Forensic Psychologists) and mental health consumer and carer advocate organisations (e.g., National Mental Health Consumer and Carer Forum) to disseminate through their networks.

Prospective participants expressed their interest to participate in a series of iterative focus groups via an online link in the invitation email. A selection matrix was used to select individuals to participate to ensure diversity across jurisdiction and role. Participants were asked to attend all focus group sessions if selected to participate. To mitigate power imbalance, lived experience participants were prioritised; any individuals affiliated with forensic mental health services from the same jurisdiction were not selected to participate. Focus groups were limited to 5-8 participants to enable in-depth discussion of this complex topic (Krueger and Casey, 2009).

### Data collection

A series of three semi-structured online focus groups were convened in February 2022. Questions focused on clinically relevant characteristics to define service needs and corresponding need groups in prisons (see Box 1). Questions were informed by existing NMHSPF principles and framed within the context of modelling ideal/adequate care.

#### Box 1.

*Indicative focus group questions*

- Drawing on your experiences with prison populations, and thinking of people who have a mild/moderate/severe mental illness in that setting, what are the factors that contribute to/drive an individual’s need for mental health services while they are in prison?
- Based on the factors we have just discussed, and drawing on your experiences, do you think there are any distinct sub-groups of people in prison with mild/moderate/severe mental illness who have a similar intensity and mix of mental health service needs?
- The following care profile from the NMHSPF is for people with mild/moderate/severe mental illness. Drawing on your experience, do you think this mix of services is correct for people in prisons? (display care profile on screen)

`o` Are any services missing?
`o` Are any services inappropriate?
`o` Are the proportions of the need group requiring the service appropriate?
`o` Are the providers appropriate?
`o` Is the amount of services included appropriate?

Existing need group descriptions and care profiles from across various NMHSPF severity levels (mild, moderate and severe) were used as a starting point to guide discussion about the types of characteristics used to define need groups and the types of services required for each need group in prisons (Gossip et al., 2023). NMHSPF care profiles describe the range of service types and interventions required annually on average by people in a need group, including the proportion of the group likely to require the service, who the service should be provided by and the number and duration of sessions (Diminic et al., 2023). Through an iterative co-production process across the three focus group sessions, need group descriptions and care profiles were adjusted to suit prison populations.

Focus groups were facilitated by CC with support from JJ and SD. Each session was recorded using Zoom software and transcribed verbatim with minor editing applied via an external transcription service. Transcripts were reviewed for accuracy and deidentified by the research team.

Following focus groups, all participants and stakeholders identified during recruitment were invited to complete an online Qualtrics survey to review initial need group and care profile descriptions. Survey participants rated their agreement with the information presented using four-point Likert scales and commented on the need groups and care profiles using free text boxes (Box 2).

#### Box 2.

*Online survey questions*

For each need group:

- The description of this group is appropriate (strongly disagree, disagree, agree, strongly agree) [Likert scale]
- The mix of services modelled for this group is appropriate (strongly disagree, disagree, agree, strongly agree) [Likert scale]
- Do you have any feedback about the description of the need group? [free text]
- Do you have any feedback about the mix of services or amount of care modelled in this care profile? [free text]

Overall:

- Are there any other populations who receive mental health services in prison who are not covered by these need group descriptions?
- Do you have any other comments about the prison need groups or care profiles that were not covered previously?

### Analysis

Thematic analysis of focus group transcripts and free-text survey responses was used to validate and refine initial need group and service mix descriptions. Coding was undertaken at a semantic level using a deductive framework based on non-forensic NMHSPF need group descriptions and care profiles (Table 1) (Byrne, 2022; Diminic et al., 2023). Two researchers (CC and MB) independently coded transcripts using NVivo 14 (Lumivero, 2024), with discrepancies resolved through discussion. (Byrne, 2022; Diminic et al., 2023). Additional inductive codes were developed iteratively to capture emergent themes not represented within pre-specified NMHSPF categories. Coded data were synthesised to generate final themes.

**Table 1.** Deductive thematic analysis coding scheme.

|  | **Categories** |  |
| --- | --- | --- |
|  | **Key attributes** | **Service mix** |
| **Sub-categories** | Comorbidity & Complexity<br>Psychosocial Disability<br>Risk<br>Severity | Assessment<br>Bed-based care<br>Care coordination and liaison<br>Cultural supports<br>Family and carer support<br>Peer support<br>Pharmacotherapy<br>Physical health needs<br>Psychosocial support<br>Review<br>Secure Treatment Order<br>Structured therapies<br>Transition support |

#### Analysis of Likert Scale responses

Descriptive statistics were generated for online survey responses to Likert Scales to determine the levels of agreement with survey content. A high level of agreement with survey content was defined as at least 80% of respondents answering the same way (Thomas et al., 2014).

## Results

Fourteen stakeholders from six jurisdictions expressed their interest to participate in the focus groups; ten were invited and eight attended across three sessions (Figure 2). Focus group participants were from four jurisdictions (Table 2). Three participants had a lived experience of mental illness while in prison or caring for someone with a mental illness while in prison.

Fifteen participants from five jurisdictions completed the online survey. Just over half of survey participants had a lived experience when including people employed as peer workers in prison mental health services (n=8). Five survey respondents had also participated in the prison mental health services focus groups prior to completing the online survey.

Across the focus groups and survey there were 18 unique participants. All service-related participants were affiliated with prison mental health services. There were no participants representing prison/custodial health services or NGO service partners.

**Table 2.** Participant characteristics.

|  | **Focus groups**<br>**n (%)** | **Online survey**<br>**n (%)** |
| --- | --- | --- |
| **Stakeholder type** |  |  |
| Service director (*i.e., responsible for the management and operation of a health service*) | 3 (38) | 3 (20) |
| Service manager (*i.e., employed in a health service who provide leadership, supervision and development of clinical teams*) | 1 (13) | 1 (7) |
| Service provider<sup>\$</sup> (*i.e., employed in a health service to provide care to consumers*) | 1 (13) | 7 (47) |
| Lived experience | 3 (38) | 4 (27) |
| **Jurisdiction** |  |  |
| Australian Capital Territory | 1 (13) | 1 (7) |
| New South Wales | 2 (25) | 5 (33) |
| Northern Territory | 0 (0) | 0 (0) |
| Queensland | 1 (13) | 4 (27) |
| Tasmania | 0 (0) | 0 (0) |
| South Australia | 0 (0) | 0 (0) |
| Victoria | 4 (50) | 3 (20) |
| Western Australia | 0 (0) | 2 (13) |

<sup>\$</sup> Includes peer workers

**Figure 2.**
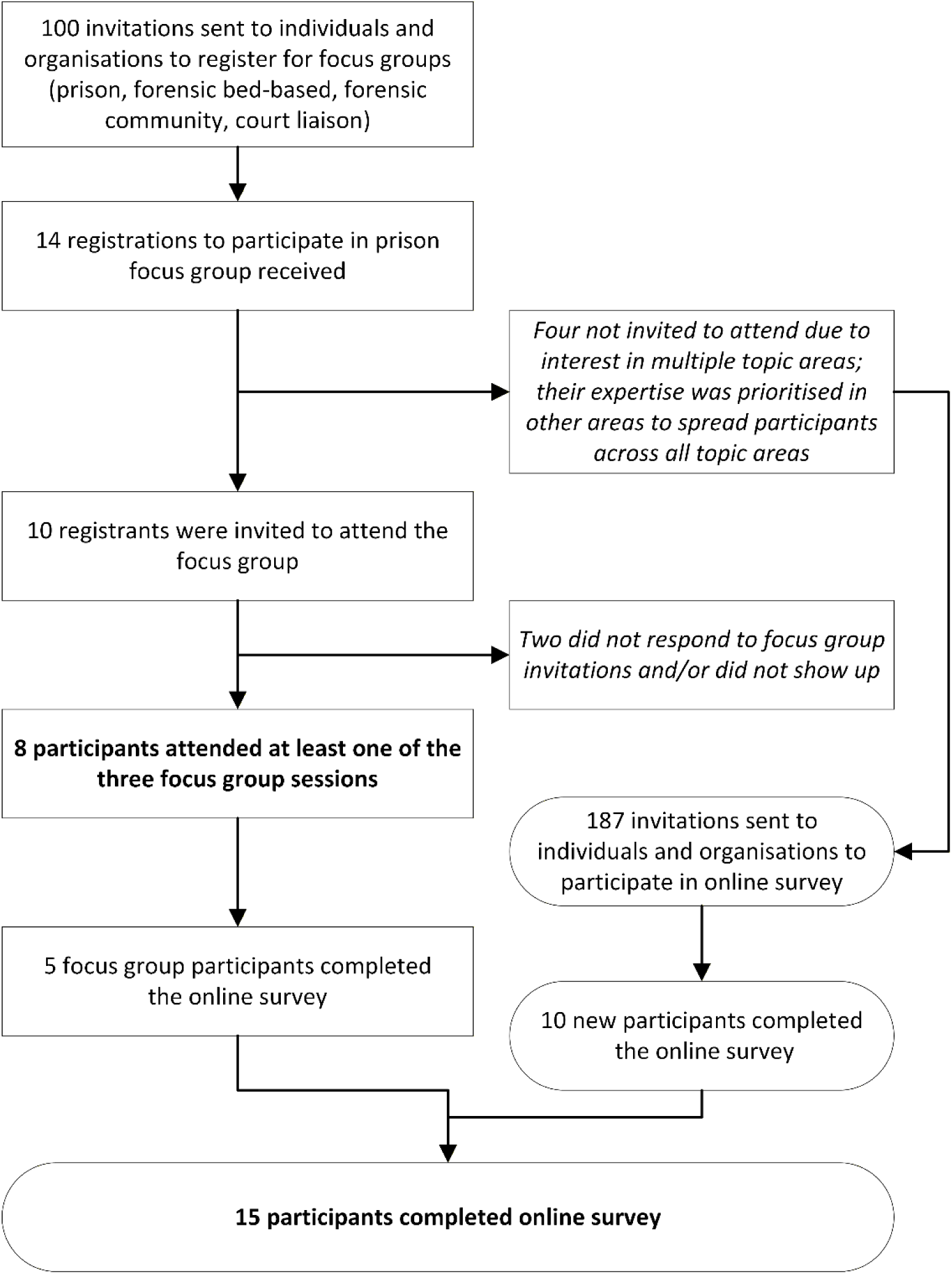
Focus group recruitment flowchart.

### Need groups identified

Four distinct need group descriptions were generated for prison populations: indicated prevention; mild; moderate; and severe and complex (see Table 3). These varied from the NMHSPF need groups initially proposed to focus group participants during the first focus group session (mild, moderate, and severe). Focus group participants identified the need for an indicated prevention need group due to the vulnerable and at-risk population with subthreshold mental illness in the prison setting. Focus group participants also advised that in the prison setting, all people with severe mental illness should be considered as severe and complex due to the high levels of comorbidity, co-occurring justice involvement, and presence of environmental stressors in prisons.

Need groups were commonly delineated by symptoms, the presence of any diagnosis, comorbidities, impact on daily functioning, and risk. All need groups experienced added social and environmental stressors related to living in a custodial setting. People in the mild, moderate, and severe and complex need groups had a diagnosed mental illness. People in the indicated prevention group had subthreshold mental illness and experience symptoms that may be related to the inherent stressors of living in a custodial setting. People in the indicated prevention group had varying needs; some may be able to function with minimal impairment while others may experience moderate to high levels of impairment in their daily lives. The impact of mental illness on daily functioning increases across the remaining severity levels, with people in the mild group able to function with minimal functional impairment, while those in the severe and complex group experience a high degree of functional impairment. The moderate and severe and complex groups experience added complexity, such as comorbid conditions. The level of risk posed by people within each need group also increases with severity, whereby the moderate and severe and complex groups may experience increased risk of harm to themselves or others.

Survey respondents had high levels of agreement (≥90%) with the identified need groups and their descriptions. People who require antilibidinal treatment were identified as an additional need group to consider by two respondents. One respondent questioned whether intellectual disability is included within the proposed need groups. Focus group participants also expressed the need to consider the distinct needs of First Nations Australians in prison settings.

**Table 3.**
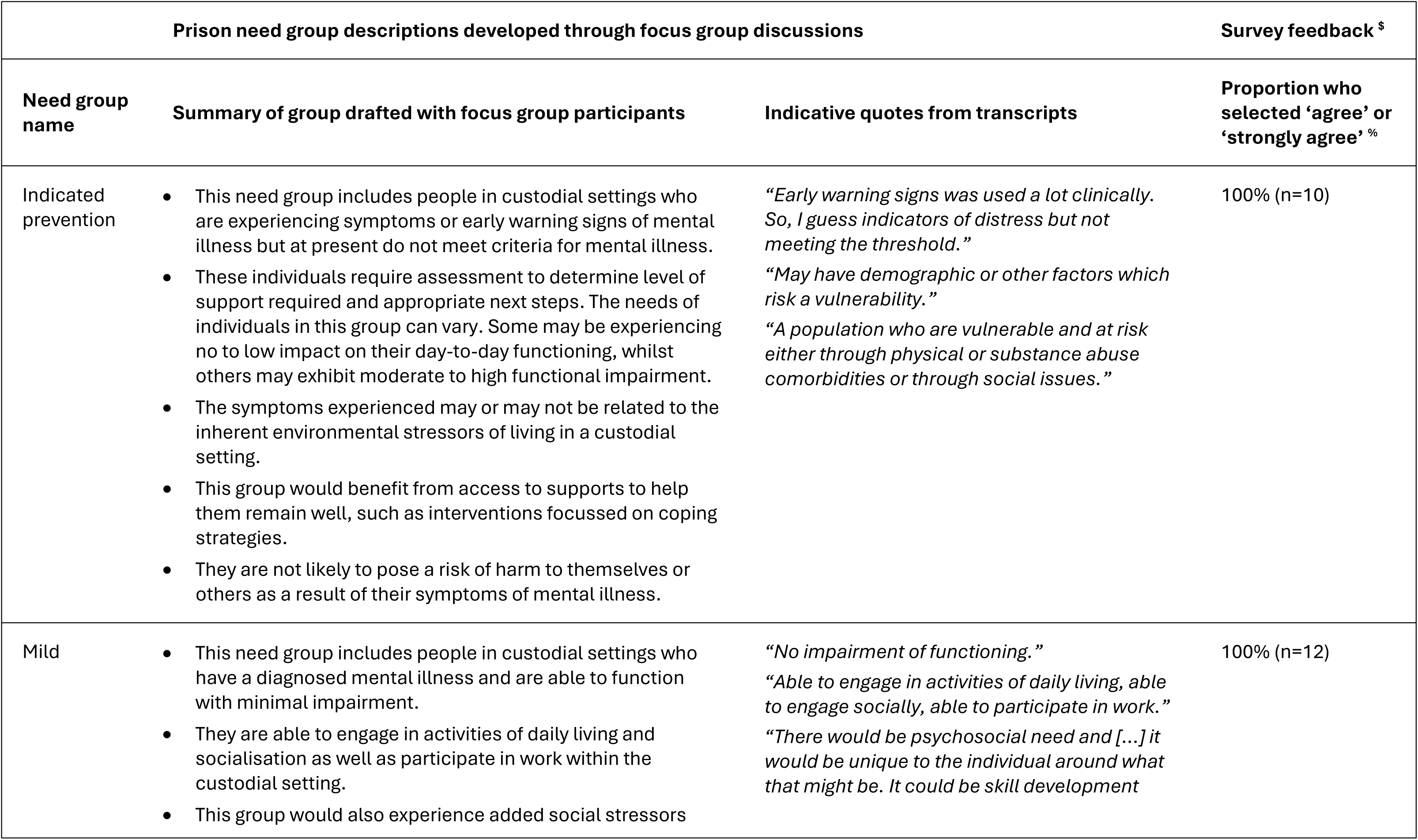
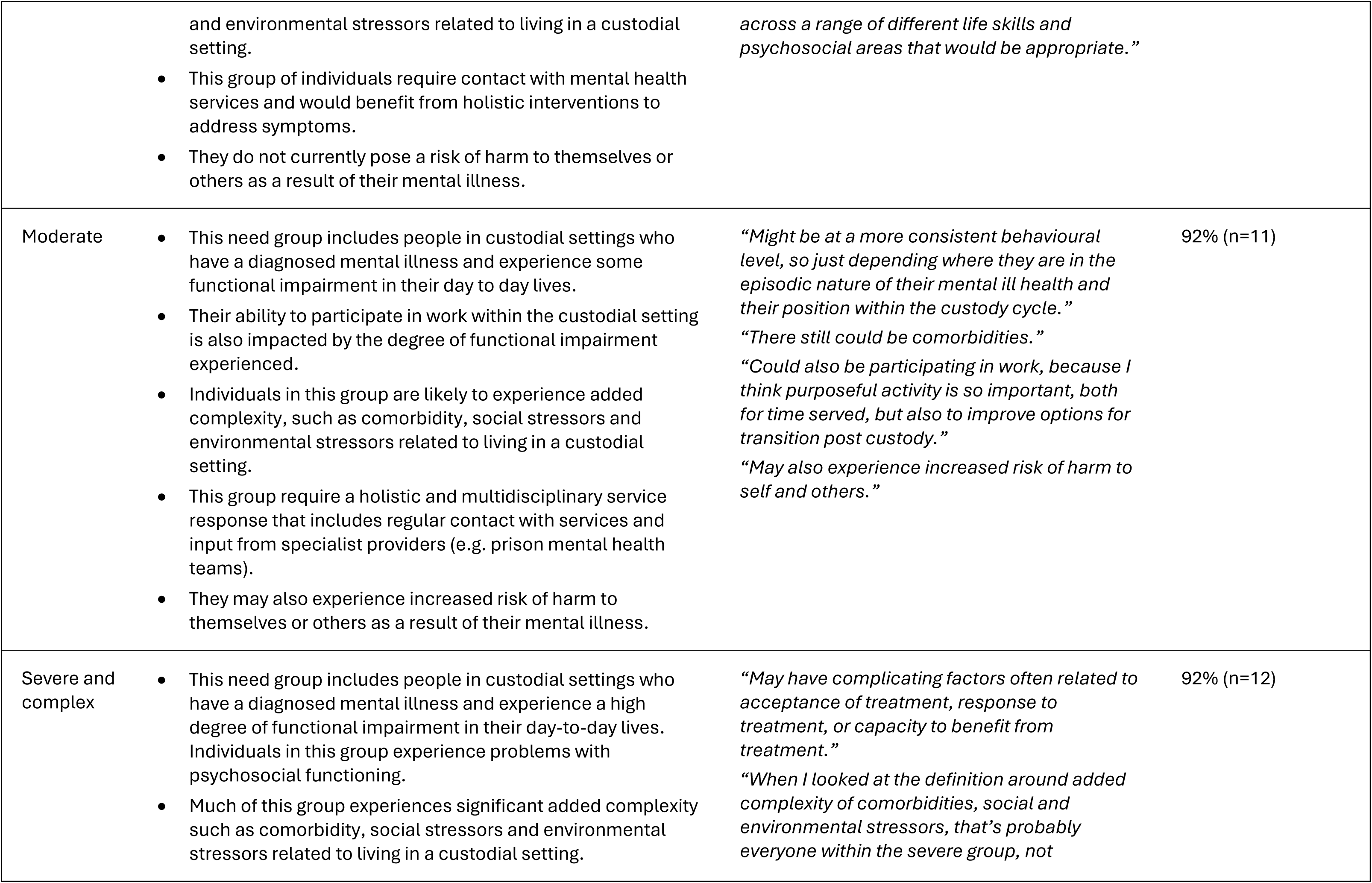
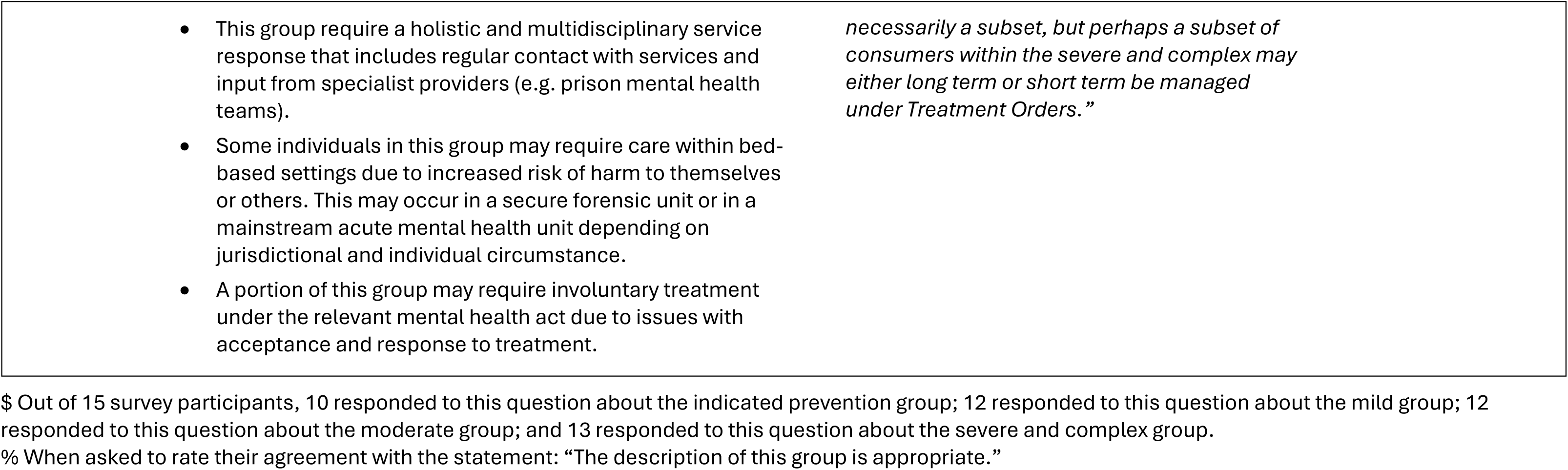
Population need groups identified and key characteristics.

### Attributes of people who require mental health care in prisons

Participants described several attributes that apply across all need groups, as illustrated by the quotes in Table 4. First, participants reiterated that people in prisons experience greater severity of mental illness than people with mental illness in the community and may require a higher level of support (Q4.1 and Q4.2). Participants also stated that a majority of people in prison would meet criteria for having at least a mild mental illness (Q4.3)

Comorbidity and complexity were raised as common attributes across all people requiring mental health care in prison settings. Participants stated that multimorbidity, such as alcohol and drug use, foetal alcohol spectrum disorder, and trauma are common across prison populations. Inherent social and environmental stressors associated with incarceration also increase complexity across all need groups (Q4.4)

Reception into prison and transfers between prisons and to the community upon release were raised as a particular stressor associated with higher levels of individual risk. Gender was also raised as a factor associated with greater complexity in prison settings, particularly for women whose incarceration has separated them from young children and experience associated grief and loss.

Participants also stated that the key attributes defining an individual’s need for mental health services are not static, and that people may move between groups depending on intervening stressors and other fluctuations in mental health status (Q4.5 and Q4.6).

**Table 4.**
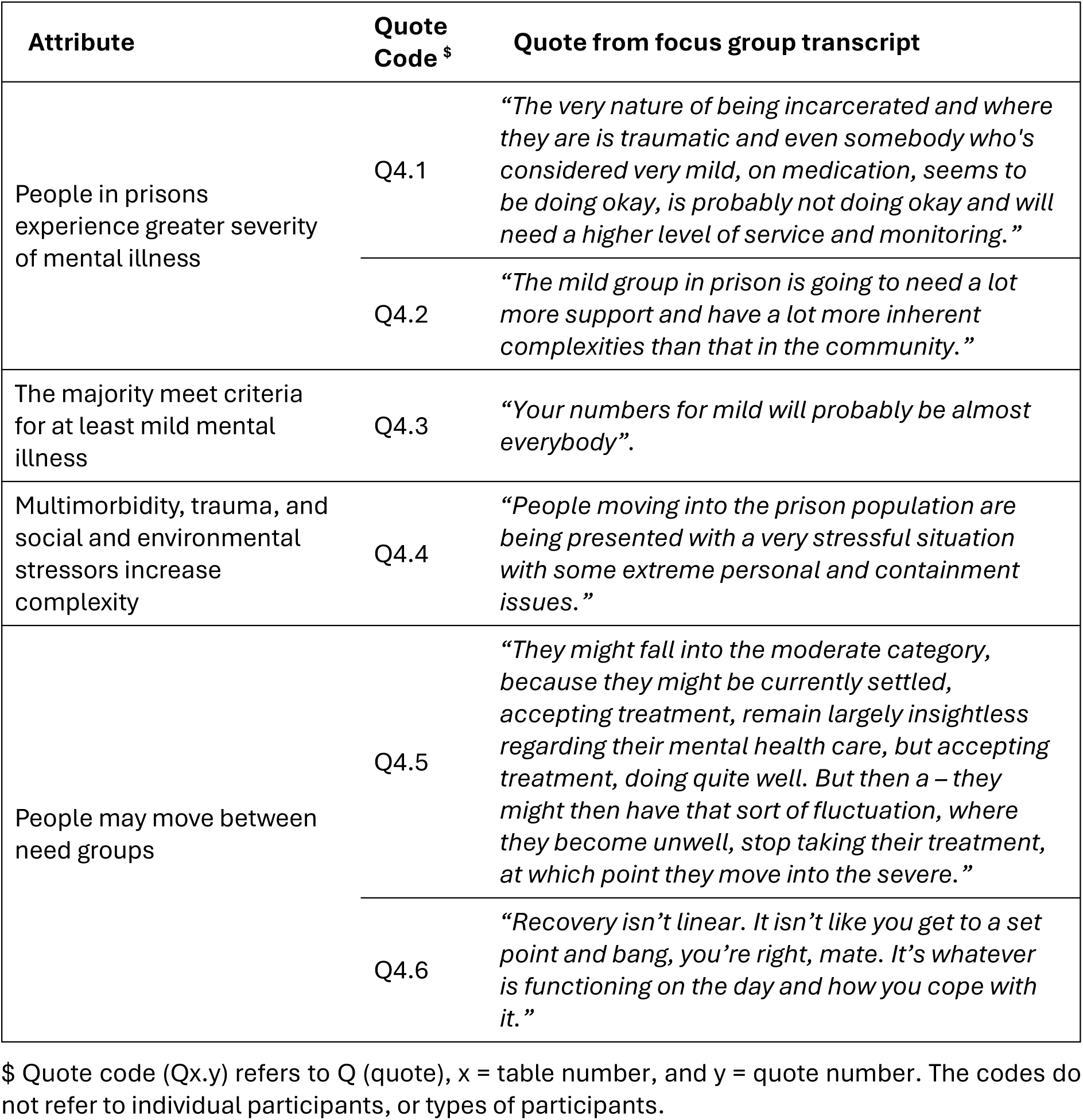
Focus group participant quotes highlighting key attributes common across all need groups.

### Mix of services required by each need group

A summary of the services required by each need group is provided in Table 5. Common service types across all need groups included assessments, psychological therapies, peer support, and lifestyle interventions. Psychosocial supports are also required across most levels of severity, except for the mild group (who are expected to experience minimal functional impairment). At the lower severity levels, various modalities may be offered for psychological therapies, including the provision of web-based programs. Comprehensiveness of assessments increases with severity level, including the need for physical health assessments and monitoring to manage physical health comorbidities. Physical health assessments still require input from specialist mental health workforce including psychiatrists and mental health nurses. These physical health assessments are relevant for the moderate and severe and complex groups largely due to the increased physical health impacts due to some psychiatric medications that may be more commonly used with greater severity of mental illness.

The relevant workforce also changes across severity levels; for example, assessments for the indicated prevention and mild levels can be provided by mental health professionals within prison primary care teams, whereas for moderate and severe and complex, these should be done exclusively by specialists within multidisciplinary prison mental health teams. Several service types are unique to the severe and complex group, including case management, care coordination with internal and external agencies, and the need for admission to acute, bed-based mental health care in a mainstream or secure setting.

Survey respondents had high levels of agreement (≥90%) with the identified service mix for each need group.

**Table 5.**
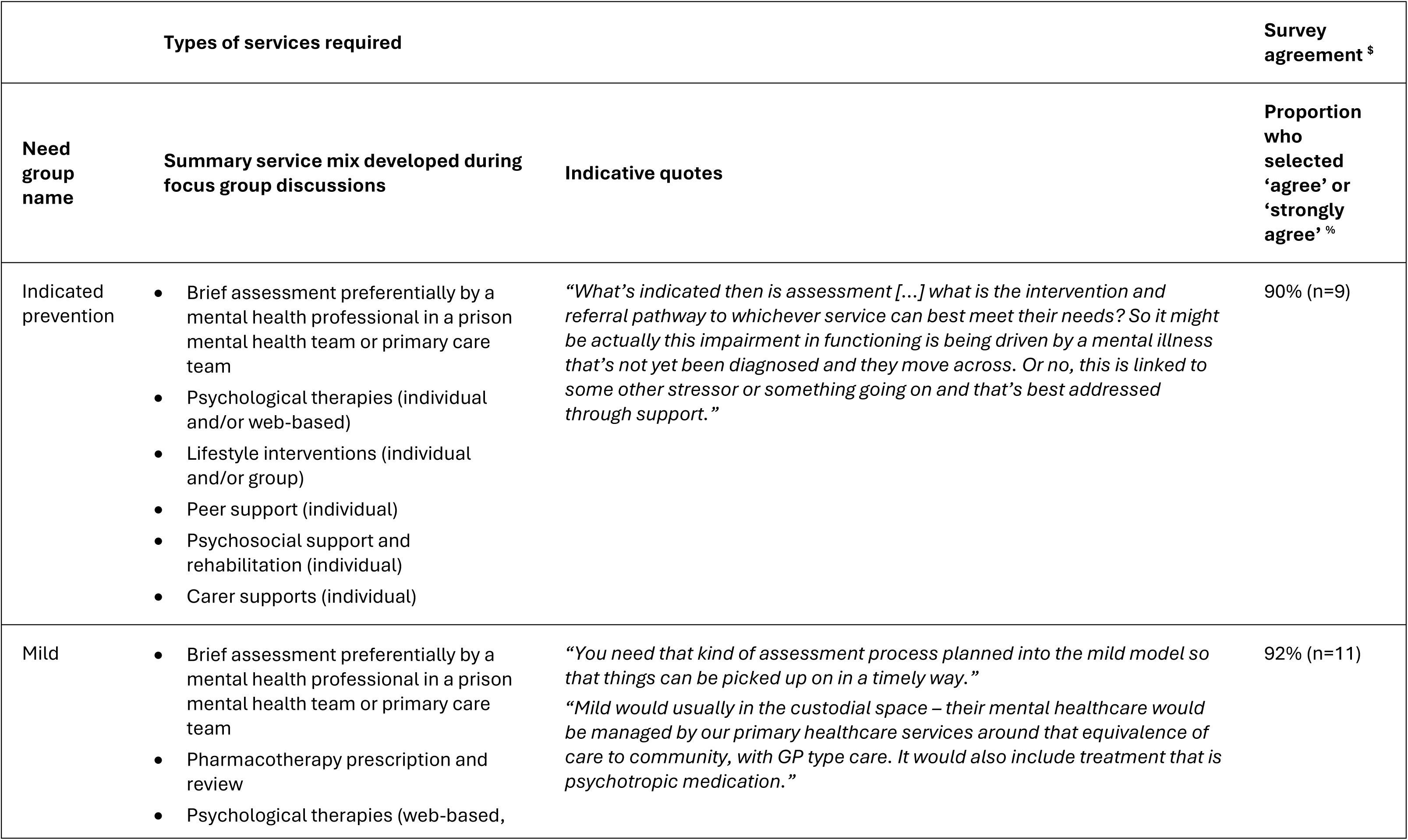
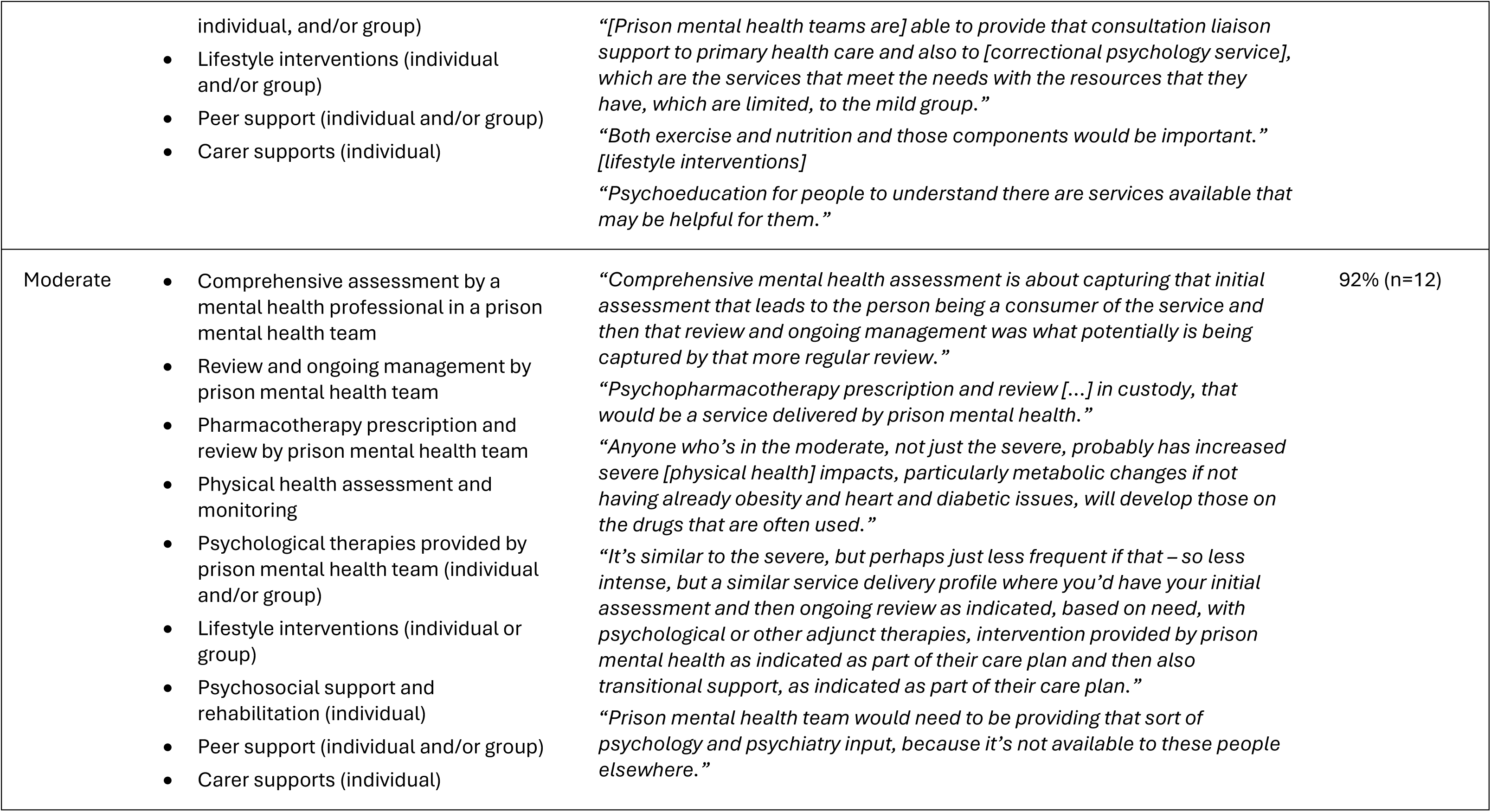
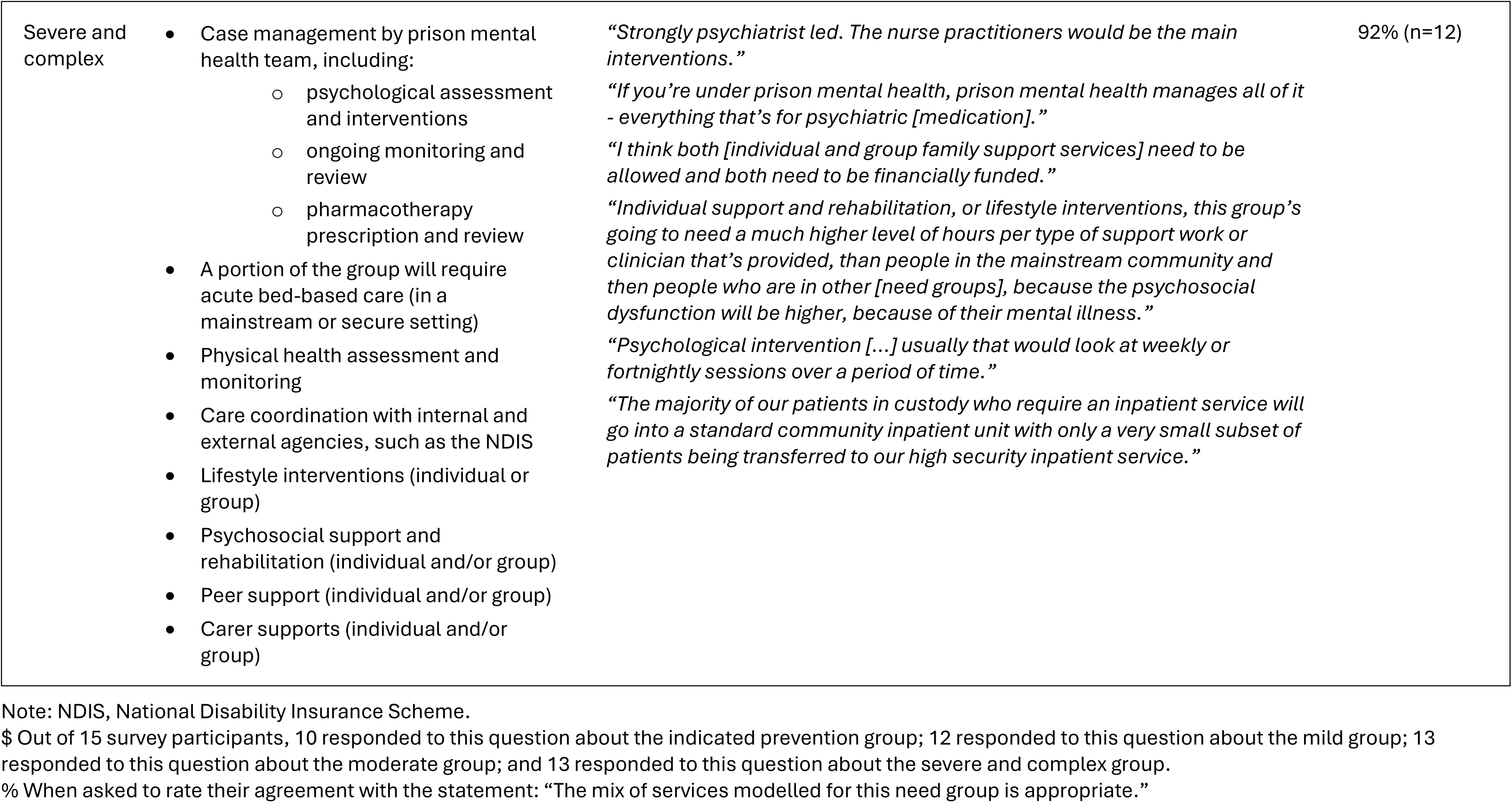
Service mix for each identified need group.

| Types of services required | | | Survey agreement <sup>\$</sup> |
| --- | --- | --- | --- |
| Need group name | Summary service mix developed during focus group discussions | Indicative quotes | Proportion who selected 'agree' or 'strongly agree' <sup>%</sup> |
| Indicated prevention | Brief assessment preferentially by a mental health professional in a prison mental health team or primary care team<br>Psychological therapies (individual and/or web-based)<br>Lifestyle interventions (individual and/or group)<br>Peer support (individual)<br>Psychosocial support and rehabilitation (individual)<br>Carer supports (individual) | *“What’s indicated then is assessment [...] what is the intervention and referral pathway to whichever service can best meet their needs? So it might be actually this impairment in functioning is being driven by a mental illness that’s not yet been diagnosed and they move across. Or no, this is linked to some other stressor or something going on and that’s best addressed through support.”* | 90% (n=9) |
| Mild | Brief assessment preferentially by a mental health professional in a prison mental health team or primary care team<br>Pharmacotherapy prescription and review<br>Psychological therapies (web-based, | *“You need that kind of assessment process planned into the mild model so that things can be picked up on in a timely way.”*<br>*“Mild would usually in the custodial space – their mental healthcare would be managed by our primary healthcare services around that equivalence of care to community, with GP type care. It would also include treatment that is psychotropic medication.”* | 92% (n=11) |
|  | individual, and/or group)<br>• Lifestyle interventions (individual and/or group)<br>• Peer support (individual and/or group)<br>• Carer supports (individual) | *“[Prison mental health teams are] able to provide that consultation liaison support to primary health care and also to [correctional psychology service], which are the services that meet the needs with the resources that they have, which are limited, to the mild group.”*<br>*“Both exercise and nutrition and those components would be important.”*<br>*[lifestyle interventions]*<br>*“Psychoeducation for people to understand there are services available that may be helpful for them.”* |  |
| Moderate | • Comprehensive assessment by a mental health professional in a prison mental health team<br>• Review and ongoing management by prison mental health team<br>• Pharmacotherapy prescription and review by prison mental health team<br>• Physical health assessment and monitoring<br>• Psychological therapies provided by prison mental health team (individual and/or group)<br>• Lifestyle interventions (individual or group)<br>• Psychosocial support and rehabilitation (individual)<br>• Peer support (individual and/or group)<br>• Carer supports (individual) | *“Comprehensive mental health assessment is about capturing that initial assessment that leads to the person being a consumer of the service and then that review and ongoing management was what potentially is being captured by that more regular review.”*<br>*“Psychopharmacotherapy prescription and review [...] in custody, that would be a service delivered by prison mental health.”*<br>*“Anyone who’s in the moderate, not just the severe, probably has increased severe [physical health] impacts, particularly metabolic changes if not having already obesity and heart and diabetic issues, will develop those on the drugs that are often used.”*<br>*“It’s similar to the severe, but perhaps just less frequent if that – so less intense, but a similar service delivery profile where you’d have your initial assessment and then ongoing review as indicated, based on need, with psychological or other adjunct therapies, intervention provided by prison mental health as indicated as part of their care plan and then also transitional support, as indicated as part of their care plan.”*<br>*“Prison mental health team would need to be providing that sort of psychology and psychiatry input, because it’s not available to these people elsewhere.”* | 92% (n=12) |
| Severe and complex | • Case management by prison mental health team, including:<br>○ psychological assessment and interventions<br>○ ongoing monitoring and review<br>○ pharmacotherapy prescription and review<br>• A portion of the group will require acute bed-based care (in a mainstream or secure setting)<br>• Physical health assessment and monitoring<br>• Care coordination with internal and external agencies, such as the NDIS<br>• Lifestyle interventions (individual or group)<br>• Psychosocial support and rehabilitation (individual and/or group)<br>• Peer support (individual and/or group)<br>• Carer supports (individual and/or group) | *“Strongly psychiatrist led. The nurse practitioners would be the main interventions.”*<br>*“If you’re under prison mental health, prison mental health manages all of it - everything that’s for psychiatric [medication].”*<br>*“I think both [individual and group family support services] need to be allowed and both need to be financially funded.”*<br>*“Individual support and rehabilitation, or lifestyle interventions, this group’s going to need a much higher level of hours per type of support work or clinician that’s provided, than people in the mainstream community and then people who are in other [need groups], because the psychosocial dysfunction will be higher, because of their mental illness.”*<br>*“Psychological intervention [...] usually that would look at weekly or fortnightly sessions over a period of time.”*<br>*“The majority of our patients in custody who require an inpatient service will go into a standard community inpatient unit with only a very small subset of patients being transferred to our high security inpatient service.”* | 92% (n=12) |

Note: NDIS, National Disability Insurance Scheme.
\$ Out of 15 survey participants, 10 responded to this question about the indicated prevention group; 12 responded to this question about the mild group; 13 responded to this question about the moderate group; and 13 responded to this question about the severe and complex group.
% When asked to rate their agreement with the statement: “The mix of services modelled for this need group is appropriate.”

### Types of services required in prisons

Across all need groups, participants described the importance of several care principles, as illustrated by select quotes from the focus group transcripts in Table 6. These included trauma-informed care; recovery-focussed care; and the principle of equivalence of care to that offered outside of prisons. Additionally, choice in seeking mental health care was considered important. Participants stated that services/service mix should not be ‘one size fits all’ and service responses should be tailored to individual needs (Q6.1).

Additionally, participants elaborated on the impact of the prison setting itself on service needs, as compared to the general community. For example, the mental health presentations of people in prison have increased complexity, resulting in a need for more intensive service responses than would be required by similar populations in the community. This may in part be due to do the lack of alternative service options available in prison that would be available to people within the community (Q6.2, Q6.3, Q6.4).

The importance of several service types across all need groups was reiterated through focus group discussions and survey feedback. First, participants described the need for universal screening upon reception into custody (Q6.5 and Q6.6), regular, ongoing mental health assessment for people in prison (Q6.7), and the need for support upon movement from one correctional facility to another (Q6.8).

It was important to participants that there was provision of intensive transition support when people in prison are preparing for release. This includes liaison with community services, including housing and the National Disability Insurance Scheme (NDIS) funding program for disability supports. Participants reiterated the need for release planning and supportive transfer of care, including linkage with relevant services in the community prior to their release (Q6.9) as a well as education and support to avoid re-incarceration (Q6.10).

Another important service type discussed across all need groups was support for the families and carers of people in prisons. Focus group participants reiterated that individual and group-based carer support services are important throughout the carer journey, including psychoeducation, support during court and tribunal processes, and transition options, including navigating the NDIS and housing options (Q6.11).

Finally, participants highlighted the importance of including peer support in the provision of mental health care in prisons. One survey participant reiterated the critical need for all care profiles to include peer workforce (Q6.12 and Q6.13).

**Table 6.** Focus group participant quotes highlighting common mental health services required across all need groups.

| **Attribute** | **Quote Code<sup>\$</sup>** | **Quote from focus group transcript** |
| --- | --- | --- |
| Service responses should be tailored to individual needs | Q6.1 | *“The mix of services will vary for different patients”.* |
| More intensive service responses are required in prisons | Q6.2 | *“Around complexities and level of service need compared to say, community space, I would say we probably have more frequent reviews in the custodial environment to what we would have in community [...] whilst we might ideally see someone more regularly, the capacity to see them more regularly is significantly impacted within the environment.”* |
|  | Q6.3 | *“[The need for] psychological therapies would be higher [within prison populations] when compared to what is offered under Medicare options in the community for individuals with less co-morbid issues and without the environmental stressors.”* |
|  | Q6.4 | *“The actual average service delivery of that, which is again going to look different to the community because, for example, we don’t have Medicare mental health care plans being provided in custody, the same way when you look at community, where you get access to all of these community services.”* |
| Universal screening is needed upon reception into custody | Q6.5 | *“Everyone who comes into custody should have a mental health screen by a mental health professional”.* |
|  | Q6.6 | *“Screening is important to identify need for onward referral to specialist [mental health] assessment.”* |
| Ongoing mental health assessments are required | Q6.7 | *“There needs to be planned into the model, consistent assessment at whatever period of time would clinically be realistic, whether it’s the one month and then three month or one month and six month.”* |
| Support is required when people are moved between correctional facilities | Q6.8 | *“Some kind of screening or assessment or follow up or opportunities for people when they’re transferred [...] they may have been doing great in one place and then been relocated or shifted from – wherever they’ve shifted to within a place might have other impacts on them.”* |
| Establish linkage with relevant community services prior to release | Q6.9 | *“It’s something that actually should be funded [...] for all of those who need it [...], because some people do have good supports before they come in to custody in the community, in there briefly and then they’re back out with those supports. But unfortunately, that’s not the majority.”* |
| Education is provided to help prevent re-incarceration | Q6.10 | *“Services need to include how to avoid re-incarceration methods and skills, and how to access relevant support services that actively support and assist this.”* |
| Support for families and carers is important | Q6.11 | *“The families [are] the ones who have to live with these people when they come back out and deal with the repercussions and the stigma and everything else that goes with whatever has put the person in the system in the first place.”* |
| Peer workforce should be included in service provision | Q6.12 | *“Utilisation of peers in all areas of interaction between service providers and person needs to take place, to minimise the stigma associated with being incarcerated while having mental health concerns or barriers.”* |
|  | Q6.13 | *“Group based consumer peer support and individualised peer support have a much larger role both in [this] setting and on release”.* |

\$ Quote code (Qx.y) refers to Q (quote), x = table number, and y = quote number. The codes do not refer to individual participants, or types of participants.

## Discussion

This study defined the characteristics and service needs of four distinct population groups in prisons who require mental health care. These groups were delineated using characteristics including presence of a diagnosed mental illness, level of functional impairment, presence of added complexity, and service response required. The need groups and key services were identified by a broad range of prison mental health stakeholders across Australia, including lived experience representation.

People in prison are entitled to health care equivalent to that available in the community across the entire continuum of care, and across all levels of need (Lines, 2006) (Bower and Gilbody, 2005; Perkins, 2016). A key finding of this study is the identification of a need group comprising people in prison who do not meet diagnostic thresholds, but who display risks or vulnerabilities of developing mental health conditions. This is an important inclusion, as it allows the provision of preventative care prior to the emergence of a diagnosable mental illness.

Across all need groups, key service types included universal screening on reception, routine assessment, and intensive transition support prior to release. These components are consistent with previous research describing key elements of mental health care in prisons (Forrester et al., 2018; Ogloff, 2002) and recommendations from the recent Productivity Commission inquiry into Mental Health (Productivity Commission, 2020). However, peer support, lifestyle interventions, psychosocial supports, and support for families and carers emerged as additional elements of the service mix in prisons. There is promising evidence suggesting that peer supports are acceptable to people in prison and have positive effects (Bagnall et al., 2015), including in supporting reintegration upon release (Treitler et al., 2025). Evidence for multidisciplinary lifestyle interventions has also increased in recent years, leading to practice and policy level recommendations for their inclusion in mental health care (Gossip et al., 2025; Manger, 2019; Productivity Commission, 2020).

Supports offered to families and carers of people with mental illness in the community typically include psychoeducation, support groups and practical assistance (Gordon et al., 2022). This support should be provided to individuals who have a family member with mental illness in prisons, including information about visitation schedules, emotional support, and additional support in anticipation of their family member being released from prison (Scottish Government, 2025).

There is a complex interplay of mental health service providers in prisons, including primary care services provided by custodial health services and prison mental health services. To enable equivalence of care, it was preferred that the mental health care for the indicated prevention and mild need groups be managed by primary care services, whereas care for the moderate and severe and complex groups should be managed by prison mental health teams. However, prison mental health teams may provide some services, including screening and assessments, to people with less severe mental illness, to ensure they are provided by a mental health professional.

Although psychological therapies were included in the service mix across all need groups, there is currently limited access in prisons (Plueckhahn et al., 2015). Clinician moderated, web-based psychological therapies were included in the service mix for the indicated prevention and mild groups, which serve as an effective alternative to face-to-face care, especially in limited resource settings (Titov et al.). In recent years there has been greater focus on increasing access to technology in prison, which would enhance the utility for these alternative service models (Franich and Martinovic, 2024).

Participants reiterated the need to understand the characteristics and mental health service mix required for high priority groups in prisons, including First Nations Australians. The need groups and service mixes described in this study do not account for the specific cultural needs of First Nations Australians; this is an important next step. Other need groups were also suggested, including people who require anti-libidinal treatments and people with intellectual disability. The need groups and service mixes described here are for people with mental illness or mental health distress but are not exclusionary to people who may experience comorbidities or additional service needs.

### Limitations

Understanding the drivers of mental health service needs in prisons is limited by the challenges of conducting research in correctional settings. These challenges include the complexities of research and ethical approvals, the need for additional resourcing for security staff during research activities, and the difficulties of designing survey instruments that account for literacy, comprehension, and attentional challenges (Gaber et al., 2025). Critically, prison populations are excluded from other information sources that would help to generate an understanding of the characteristics driving service needs of people with mental illness in prisons, such as Australia’s National Study of Mental Health and Wellbeing (Australian Bureau of Statistics, 2022a). These considerations reiterate the importance of using expert clinical guidance to help define the populations requiring mental health care in prisons.

Second, this study encompassed a small sample drawn from only four jurisdictions, and did not include some invited stakeholder groups, such as custodial health services and non-government organisations providing transitional support. Participants were from Australia’s most populous states and territories (Queensland, New South Wales, Victoria and Australian Capital Territory) and may have provided different insights than unrepresented jurisdictions, which have differing legislation and service delivery challenges (David McGrath Consulting, 2019; Heffernan et al., 2015; Tasmanian Government, 2019). The online survey aimed to include states and territories that were not represented in the focus groups and ultimately received a response from one additional jurisdiction.

Third, there was limited lived experience involvement in this study. This limitation is important to note, as people with a lived experience of using mental health services in prisons may have had different perspectives on the mix of mental health services required in prisons. Although the sampling matrix used to assign participants to focus groups prioritised lived experience participants, only a small number of people with a lived experience of using prison mental health services signed up to participate in the study.

### Implications

The prison-specific need groups are multidimensional, encompassing mental health, physical health, environmental health, and social health. This sets an important foundation for matching the types and intensity of treatment required to meet service needs across these dimensions of health (Sawrikar et al., 2021). Additionally, the multifactorial approach used in this study was able to expand on previous studies that used single-factor indicators such as diagnosis (Crocker et al., 2015), offence type (Adams et al., 2018) and gender (Stewart et al., 2021) to describe mental health service needs in forensic mental health settings. The information generated through this study forms part of the evidence required to develop a needs-based forensic mental health planning model in Australia. The creation of such a model will help service planners to identify gaps and generate resourcing targets to ensure that mental health services in prisons are well-resourced and available to all who need them, ultimately helping to improve service access. The diverse mix of services and providers identified for each need group will require effective cooperation between the various stakeholders involved in provision of mental health care in prisons.

## Data Availability

The data that support the findings of this study are available upon reasonable request from the corresponding author. The data are not publicly available due to privacy or ethical restrictions.

## Statements and declarations

### Ethical considerations

Ethics approval was obtained from the UQ Human Research Ethics Committee (2021/HE002377).

## References

Adams J, Thomas SDM, Mackinnon T, et al. (2018) The risks, needs and stages of recovery of a complete forensic patient cohort in an Australian state. BMC Psychiatry 18(35).

Australian Bureau of Statistics (2022a) National Study of Mental Health and Wellbeing methodology. Canberra: ABS.

Australian Bureau of Statistics (2022b) National Study of Mental Health and Wellbeing: Summary Results, 2020-21. Canberra: ABS.

Australian Government Department of Health (2020) National PHN Guidance: Initial assessment and referral for mental healthcare. Canberra.

Bagnall A, South J, Hulme C, et al. (2015) A systematic review of the effectiveness and cost-effectiveness of peer education and peer support in prisons. BMC Public Health 15(290).

Bartels L, Fitzgerald R and Freiberg A (2018) Public opinion on sentencing and parole in Australia. Probation Journal 65(3): 269–284.

Bower P and Gilbody S (2005) Stepped care in psychological therapies: access, effectiveness and efficiency. Narrative literature review. British Journal of Psychiatry 186(1): 11–17.

Butler T, Allnutt S and Yang B (2007) Mentally ill prisoners in Australia have poor physical health. International Journal of Prisoner Health 3(2): 99–110.

Butler T, Indig D, Allnutt S, et al. (2011) Co-occurring mental illness and substance use disorder among Australian prisoners. Drug and Alcohol Review 30(2): 188–194.

Byrne D (2022) A worked example of Braun and Clarke’s approach to reflexive thematic analysis. Quality & Quantity 56: 1391–1412.

Comben C, Meurk C, Rutherford Z, et al. (2026) Systematic review of available data and gaps on the prevalence of adults who require forensic mental health care in Australia. *Psychiatry*, Psychology and Law. DOI: 10.1080/13218719.2026.2624848.

Crocker AG, Nicholls TL, Seto MC, et al. (2015) The national trajectory project of individuals found not criminally responsible on account of mental disorder in Canada. Part 2: the people behind the label. The Canadian Journal of Psychiatry 60(3): 106–116.

David McGrath Consulting (2019) Report on the review of Forensic Mental Health and Disability Services within the Northern Territory.

Diminic S, Gossip K, Page I, et al. (2023) Introduction to the National Mental Health Service Planning Framework. Commissioned by the Australian Government Department of Health and Aged Care. Brisbane: The University of Queensland.

Egeressy A, Butler T and Hunter M (2009) ‘Traumatisers or traumatised’: trauma experiences and personality characteristics of Australian prisoners. International Journal of Prisoner Health 5(4): 212–222.

Forrester A, Till A, Simpson A, et al. (2018) Mental illness and the provision of mental health services in prisons. British Medical Bulletin 127(1): 101–109.

Franich G and Martinovic M (2024) Deployment of digital devices in prisons in New South Wales: exploring the benefits, challenges and opportunities for incarcerated women. Feminist Criminology 19(4): 312–328.

Gaber J, Kerrigan C, Grenada IM, et al. (2025) Continuing the conversation: practical strategies to enable acceptable and feasible health research in prisons. Archives of Public Health 83(137).

Gilling McIntosh L, Rees C, Kelly C, et al. (2023) Understanding the mental health needs of Scotland’s prison population: a health needs assessment. Frontiers in Psychiatry 14: 1119228.

Goddard T and Pooley JA (2019) The impact of childhood abuse on adult male prisoners: a systematic review. Journal of Police and Criminal Psychology 34: 215–230.

Gordon R, Grootemaat P, Loggie C, et al. (2022) Evaluation of NSW Family and Carer Mental Health Program: Final Report. Centre for Health Service Development, Australian Health Services Research Institute, University of Wollongong.

Gossip K, Diminic S, Comben C, et al. (2023) National Mental Health Service Planning Framework - Care Profiles and Top-ups - Commissioned by the Australian Government Department of Health and Aged Care. Version AUS V4.3. Brisbane.

Gossip K, John J, Comben C, et al. (2025) Estimating population-based need for lifestyle interventions among young adults with mental disorders in Australia. International Journal of Mental Health Nursing 34(2): e70034.

Heffernan E, Clugston B and Patchett S (2015) Review of the South Australian Forensic Mental Health Service. July 2015.

Holmwood C, Marriott M and Humeniuk R (2008) Substance use patterns in newly admitted male and female South Australian prisoners using the WHOASSIST (Alcohol, Smoking and Substance Involvement Screening Test). International Journal of Prisoner Health 4(4): 198–207.

Kerslake M, Simpson M, Richmond R, et al. (2020) Risky alcohol consumption prior to incarceration: a cross-sectional study of drinking patterns among Australian prison entrants. Drug and Alcohol Review 39(6): 694–703.

Krueger RA and Casey MA (2009) Focus groups: a practical guide for applied research. USA: SAGE Publications.

Lines R (2006) From equivalence of standards to equivalence of objectives: the entitlement of prisoners to health care standards higher than those outside prisons. International Journal of Prison Health 2(4): 269–280.

Linnane D, McNamara D and Toohey L (2023) Ensuring universal access: the case for Medicare in prison. Alternative Law Journal 48(2): 102–109.

Long JS, Sullivan C, Wooldredge J, et al. (2018) Matching needs to services: prison treatment program allocations. Criminal Justice and Behavior 46(5): 674–696.

Lumivero (2024) NVivo (Version 14).

Manger S (2019) Lifestyle interventions for mental health. Australian Journal of General Practice 48(10).

O’Cathain A, Murphy E and Nicholl J (2017) The quality of mixed methods studies in health services research. Journal of Health Services Research & Policy 13(2): 92–98.

Ogloff JRP (2002) Identifying and accommodating the needs of mentally ill people in gaols and prisons. *Psychiatry*, Psychology and Law 9(1): 1–33.

Patel V, Saxena S, Lund C, et al. (2023) Transforming mental health systems globally: principles and policy recommendations. The Lancet 402(10402): 655–656.

Perkins D (2016) Stepped care, system architecture and mental health services in Australia. International Journal of Integrated Care 16: 16.

Plueckhahn TM, Kinner SA, Sutherland G, et al. (2015) Are some more equal than others? Challenging the basis for prisoners’ exclusion from Medicare. Medical Journal of Australia 203: 359–361.

Productivity Commission (2020) Mental Health, Report no. 95. Canberra.

Productivity Commission (2021) Australia’s prison dilemma. Research paper. Canberra.

Richmond RL, Indig D, Butler TG, et al. (2013) Smoking and other drug characteristics of Aboriginal and non-Aboriginal prisoners in Australia. Journal of Addiction 2013(1): 516342.

Sawrikar V, Stewart E, Lamonica HM, et al. (2021) Using staged care to provide “Right Care First Time” to people with common affective disorders. Psychiatric Services 72(6): 691–703.

Scottish Government (2025) Literature review: understanding family support needs of people in prison custody.

Segal L, Guy S, Leach M, et al. (2018) A needs-based workforce model to deliver tertiary-level community mental health care for distressed infants, children, and adolescents in South Australia: a mixed-methods study. The Lancet Public Health 3(6): e296–e303.

Stewart A, Ogilvie JM, Thompson C, et al. (2021) Lifetime prevalence of mental illness and incarceration: an analysis by gender and Indigenous status. Australian Journal of Social Issues 56(2): 244–268.

Suetani S, Gill N and Salvador-Carulla L (2024) The mental health crisis needs more than increased investment in the mental health system. Medical Journal of Australia 220(9): 443–444.

Tasmanian Government (2019) Health Workforce 2040. Hobart: Department of Health.

Thomas SL, Wakerman J and Humphreys JS (2014) What core primary care services should be available to Australians living in rural and remote communities? BMC Family Practice 15(143).

Titov N, Dear BF, Staples LG, et al. (2017) The first 30 months of the MindSpot Clinic: evaluation of a national e-mental health service against project objectives. Australian & New Zealand Journal of Psychiatry 51(12): 1227–1239.

Treitler P, DiGioia-Laird V and Long B (2025) Peer support services for individuals with health-related needs reentering the community after incarceration: a scoping review of program elements and outcomes. Health & Justice 13(51).

Trofimovs J, Dowse L, Srasuebkul P, et al. (2021) Using linked administrative data to determine the prevalence of intellectual disability in adult prison in New South Wales, Australia. Journal of Intellectual Disability Research 65(6): 589–600.

World Health Organisation (2003) Planning and budgeting to deliver services for mental health (Mental Health Policy and Service Guidance Package). Geneva.

Yee N, Browne C, Rahman F, et al. (2024) Prevalence of mental illness among Australian and New Zealand people in prison: a systematic review and meta-analysis of studies published over five decades. Australian & New Zealand Journal of Psychiatry 58(12).

